# Selective effects of dopaminergic and noradrenergic degeneration on cognition in Parkinson’s disease

**DOI:** 10.1101/2024.09.16.24313753

**Authors:** Sophie Sun, Victoria Madge, Jelena Djordjevic, Jean-François Gagnon, D Louis Collins, Alain Dagher, Madeleine Sharp

## Abstract

The substantia nigra and locus coeruleus are among the first brain regions to degenerate in Parkinson’s disease. This has important implications for early cognitive deficits as these nuclei are sources of ascending neuromodulators (i.e., dopamine and noradrenaline) that support various cognitive functions like learning, memory, and executive function. However, because most studies of the relationship between patterns of degeneration and cognition have either studied these neuromodulator systems in isolation or studied specific cognitive domains in isolation, it is unknown if degeneration in the substantia nigra and degeneration in the locus coeruleus independently and selectively contribute to different cognitive deficits in Parkinson’s disease.

To address this gap, we tested people with Parkinson’s disease and older adults on tasks of positive reinforcement learning, attention/working memory, executive function, and memory to measure performance in domains of cognition specifically thought to be related to dopaminergic and noradrenergic function. Participants also underwent neuromelanin-sensitive magnetic resonance imaging which provides a measure of degeneration of dopamine neurons in the substantia nigra and of noradrenergic neurons in the locus coeruleus. Brain-behaviour relationships were evaluated by separate linear regressions predicting cognitive performance in each domain from substantia nigra and locus coeruleus neuromelanin signal intensities controlling for age, sex, and education.

As expected, Parkinson’s disease patients had significantly slower learning from positive feedback and lower performance on tests of attention/working memory, executive function, and memory than controls. Parkinson’s patients also had lower neuromelanin signal intensity in the substantia nigra and locus coeruleus. Examining brain-behaviour relationships, we found that reduced neuromelanin signal in the substantia nigra in Parkinson’s disease patients was independently associated with impaired positive reinforcement learning, controlling for changes in the locus coeruleus, but was not associated with other domains of cognition. In contrast, reduced neuromelanin signal in the locus coeruleus was independently associated with impairments in attention/working memory and executive function, controlling for changes in the substantia nigra, but not with reinforcement learning performance. These results show that substantia nigra degeneration and locus coeruleus degeneration independently and selectively contribute to cognitive deficits and therefore suggests that individual differences in the degree of neurodegeneration in these nuclei could explain the significant heterogeneity that exists in the cognitive and behavioural manifestations of Parkinson’s disease. These findings also highlight the potential value of leveraging known brain-behaviour relationships to develop performance-based measures of cognition that reflect underlying patterns of neurodegeneration.

## Introduction

The dopaminergic substantia nigra (SN) and noradrenergic locus coeruleus (LC) play crucial roles in modulating a wide range of cognitive processes.^6–9^ In Parkinson’s disease, degeneration of both these nuclei starts early.^10–14^ For instance, studies using magnetic resonance imaging (MRI) sensitive to neuromelanin, a by-product of catecholamine metabolism,^15,16^ have shown that neuromelanin in the SN and LC is already significantly reduced in the early stages of Parkinson’s disease.^2,17,18^ Correspondingly, impairments of the cognitive processes that depend on dopaminergic and noradrenergic signalling are also often already present in this early period.^14,19–23^ However, whether specific aspects of early cognitive dysfunction in Parkinson’s disease can be selectively attributed to degeneration in either one, or both, of these systems is less clear because they have largely been studied in isolation of one another or studies have focused on singular domains of cognitive performance rather than the various cognitive domains they are known to modulate. Meanwhile, delineating their independent contributions to cognition in Parkinson’s disease is essential for understanding the heterogeneity that exists in the cognitive phenotype of Parkinson’s disease and for developing methods to track clinical outcomes associated with early neurodegeneration.

There is a large body of work showing that pharmacological manipulations of dopamine state in Parkinson’s patients can remediate certain cognitive deficits,^20,24–34^ supporting the notion that degeneration in the SN plays an important role in the early cognitive deficits of Parkinson’s disease.^35^ Meanwhile, attempting to relate the severity of degeneration in the dopaminergic system to cognitive impairments has proven more difficult.^1,36–42^ One limitation of this work is that it has focused on standard domains of clinical neuropsychological testing (i.e., those recommended for classifying cognitive impairment in Parkinson’s disease)^43^ and may have failed to capture specific aspects of cognition known to depend on dopamine. Indeed, one of the most extensively studied dopamine-dependent cognitive processes in Parkinson’s disease that falls outside of standard neuropsychological testing is reinforcement learning.^44,45^ Reinforcement learning refers to the ability to update expectations on the basis of recent feedback (often measured as the computationally-defined learning rate). In Parkinson’s patients, reinforcement learning rate is believed to be impaired on the basis that positive feedback signals, carried by dopamine, are dampened.^30,31,46–53^ Though reinforcement learning has strong neurobiological grounding as a measure that could be used to track the cognitive consequences of SN degeneration, it has primarily been studied in small samples and in the context of dopaminergic medication manipulations, which are rarely feasible in larger scale clinical research. Whether reduced positive reinforcement learning is associated with the severity of SN degeneration in medicated Parkinson’s patients is unknown.

Another explanation for the mixed results linking SN degeneration with cognition is that there may be confounding effects of concurrent degeneration in the noradrenergic system. Indeed, degeneration of the noradrenergic LC is associated with attention, working memory, declarative memory, and executive function in Parkinson’s disease.^1,2,54–56^ There is also an important overlap in the cortical regions receiving projections from the SN and from the LC, as well as an overlap in cognitive functions reported to be sensitive to dopaminergic and noradrenergic modulation.^57^ Therefore, establishing a clear account of the independent effects of SN and LC degeneration on cognition will require consideration of these possible confounding effects.

Age-related brain changes also overlap with those of Parkinson’s disease. In particular, loss of LC integrity occurs with aging and is associated with cognitive impairments in older adults,^58^ yet only two studies examining LC integrity and its relationship to cognition in Parkinson’s disease accounted for the effects of age.^1,2^ It therefore remains unclear to what degree the relationship between LC degeneration and cognition observed in Parkinson’s patients is truly specific to the neurodegeneration of Parkinson’s disease.

The objective of the present study was to determine whether SN and LC neurodegeneration in Parkinson’s disease are independently associated with specific cognitive deficits. In a large sample of people with Parkinson’s disease and older adults we measured cognitive processes known to be related to these nuclei: positive reinforcement learning for the SN, and executive function, attention/working memory, and declarative memory for the LC. A subset of these participants also underwent neuromelanin MRI to extract measures of SN and LC neurodegeneration. First, we predicted that people with Parkinson’s disease would show impairments across all these measures compared to the older adults. Second, we predicted that SN neurodegeneration would be selectively associated with impaired learning from positive feedback while controlling for effects of LC degeneration and of aging and, conversely, that LC degeneration would be selectively associated with impairments in attention/working memory, executive function, and declarative memory while controlling for effects of SN degeneration and of aging.

## Methods

### Participants

One hundred and thirty-five people with Parkinson’s disease (PD) and 72 older adults (Controls) were recruited from the Quebec Parkinson Network (QPN) – a registry of patients with Parkinson’s disease who have expressed interest in research and who are referred to the network by their neurologist – or from the Montreal community in the case of older adults. Other than age (ranging from 50-90), there were no exclusion criteria to ensure a representative sample. The sample characteristics are summarized in **Table 1**. Patients completed the study assessments in their usual dopaminergic medicated “ON” state (only three patients were not taking any dopaminergic medication at the time of testing), but due to an error in data collection, the details of their dopaminergic medications are not available. All participants completed the study in their language of preference (English or French) at the Montreal Neurological Institute, provided written informed consent, and were paid $25 for their participation. The study was approved by the McGill University Health Centre Research Ethics Board.

**Table 1.** Sample Characteristics.

|  | Control<br>(n=72) | Parkinson’s disease<br>(n=135) | p-value |
| --- | --- | --- | --- |
| Age | 63.3 (10.1) | 64.3 (9.2) | 0.48<sup>a</sup> |
| Male/Female | 25/47 | 95/40 | <0.001<sup>b</sup> |
| Education, years | 15.4 (3.3) | 15.2 (3.1) | 0.56<sup>a</sup> |
| MoCA | 27.1 (1.8) | 25.8 (3.0) | 0.03<sup>c</sup> |
| Disease Duration |  | 4.9 (4.1) |  |
| UPDRS-III |  | 28.2 (13.6) |  |
| Questionnaire for Impulsive-Compulsive Disorders in Parkinson's disease | 15.0 (10.7) | 17.1 (10.6) | 0.19<sup>c</sup> |
| Geriatric Depression Scale | 1.7 (3.0) | 3.9 (4.1) | <0.001<sup>c</sup> |

Values depict mean and standard deviation.
Abbreviations: Unified Parkinson's Disease Rating Scale (UPDRS)
<sup>a</sup>Group comparison evaluated using parametric independent samples t-test
<sup>b</sup>Group comparison evaluated using chi-square test
<sup>c</sup>Group comparison evaluated using non-parametric Mann-Whitney U test

Participants completed questionnaires about demographic information and mood (Questionnaire for Impulsive-Compulsive Disorders in Parkinson’s disease and Geriatric Depression Scale), MRI, standard neuropsychological testing, and additional neurocognitive testing. Patients with Parkinson’s disease completed the Unified Parkinson’s Disease Rating Scale. Each element of the protocol is described in further detail below. The testing took place over one or two days within 90 days of each other, depending on scheduling. Due to the toll of the testing protocol, not all participants completed all assessments and therefore some of the analyses were conducted on different but largely overlapping sub-samples to maximize sample sizes and statistical power for each set of analyses. The exact sample size for each analysis is indicated in the relevant results section and the participant characteristics for each subsample are summarized in **Supplementary Table 1**. Importantly, the sample characteristics were similar across subsamples and the key group differences observed in the overall sample (for sex distribution, MoCA and depression score) were also observed in each subsample.

### Cognitive testing

#### Cognitive measure of SN degeneration

Reinforcement learning performance was measured using an adapted probabilistic learning task^48,59^ with a focus on the learning phase of the task given our interest in computationally modeling the reinforcement learning rate (**Fig. 1**). The reinforcement learning task was administered using PsychoPy version 2020.2.10 (Open Science Tools Ltd) ^60^. In the learning phase, three abstract visual stimulus pairs (AB, CD, EF) were presented, and participants learned to choose one of the two stimuli in the pair based on the feedback provided after each trial. Each pair was presented 50 times for a total of 150 trials. The order of stimulus pair presentation was randomized, as was the side of the screen on which each stimulus of a pair was presented. Feedback was probabilistic such that for the AB pair, the probability of receiving positive feedback for choosing stimulus A was 0.8, the probability of stimulus C being rewarded in the CD pair was 0.7, and the probability of stimulus E being rewarded in the EF pair was 0.6. Stimulus images were randomly assigned to each probability pair for each participant. Written feedback was presented on screen after each trial and consisted of “Correct!” or “Incorrect!”. To enhance engagement, participants were provided a task narrative: they were told that the visual stimuli represented store logos and that their goal was to accumulate as many items as possible by learning which stores were most likely to reveal the item category of interest. Participants were randomly assigned to one of two task versions: shopping for food items or shopping for household items. To support the task narrative, at the time of feedback, images of either food or household items^59,61^ were shown depending on whether the trial was rewarded or not. On trials where a participant was not rewarded, an image from the alternate category was shown.

**Figure 1.**
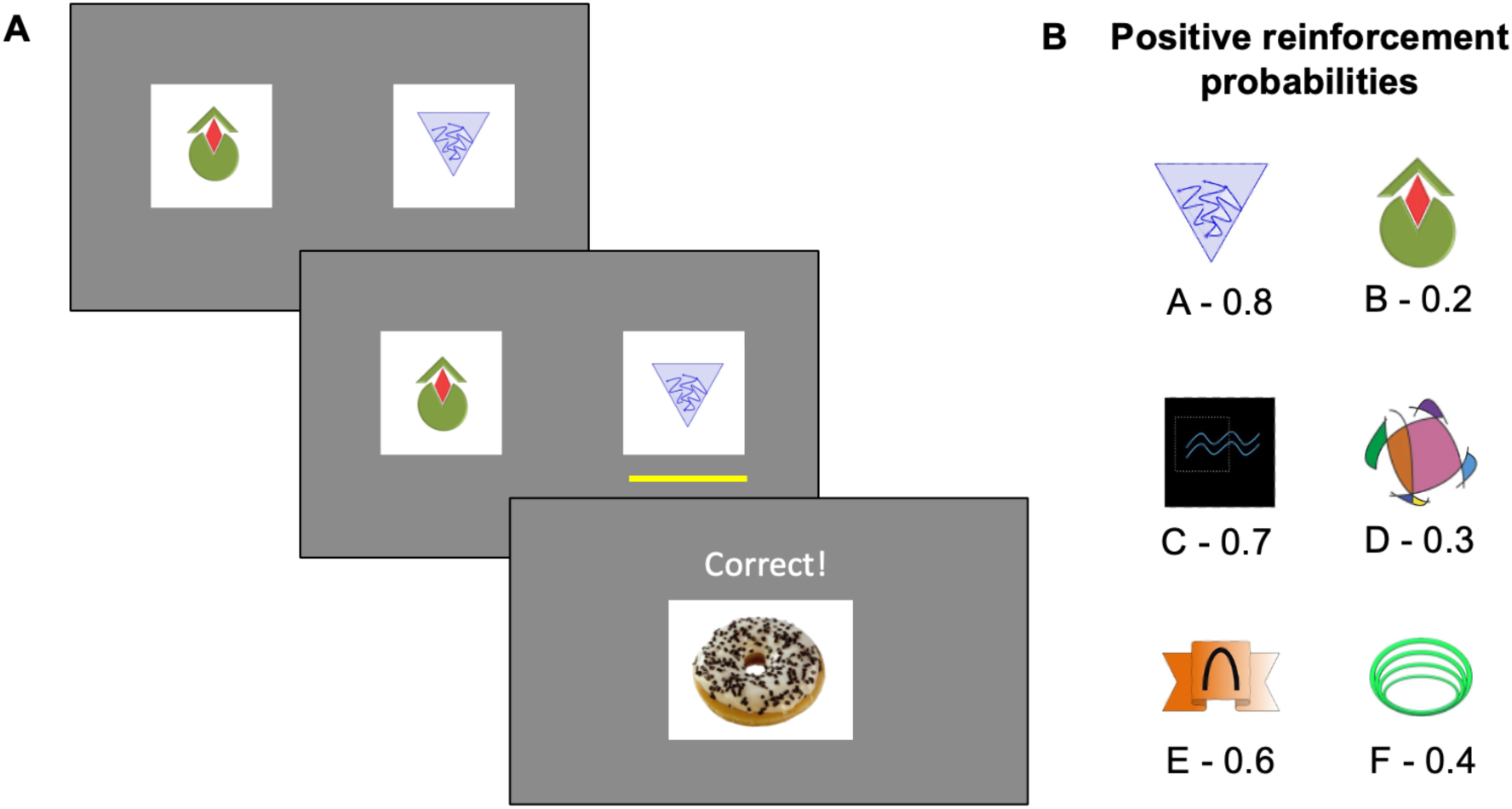
Reinforcement learning task. Participants were instructed to shop for items in a given category (i.e., food or household items) by learning which stores would most likely yield a target item through trial-and-error. **(A)** On each trial, participants had to choose between two visual stimuli, which represented the store logos. Upon selection of one of the stimuli, participants were provided with trial-unique visual feedback of a target item if they were correct or a non-target item if they were incorrect. **(B)** The stimuli had varying probabilities of yielding a correct target item with a range between 0.2 and 0.8. Shown is an example of stimulus pairs with associated probabilities.

#### Cognitive measures of LC degeneration

The measures of interest related to LC degeneration were extracted from the neuropsychological testing that participants completed. This testing protocol was designed to align with the Movement Disorder Society recommendations for assessment of cognition in Parkinson’s disease and tested five domains,^43,62^ but, because of their previously demonstrated relationship to LC, the domains of interest here were: attention/working memory, executive function and memory.^12,39,60–62^ Attention/working memory were assessed using the Digit Span test (forward and backward) and the Trail Making Test Part A (TMT A). Executive function was assessed using the TMT Part A subtracted from TMT Part B (TMT B-A), the Delis-Kaplan Executive Function (D-KEFS) Color-Word Interference Test (CWIT) using response time on the interreference condition (condition 3: Inhibition), and the Brixton Spatial Anticipation Test (BSAT; number of errors). Memory was assessed using the Hopkins Verbal Learning Test (HVLT; total on trials 1, 2, 3 and delayed recall) and the Rey Complex Figure Test (RCFT; immediate and delayed recalls). To standardize the direction of the performance scores such that greater scores represent better performance, TMT A, TMT B-A, D-KEFS CWIT, and BSAT scores were multiplied by -1.

#### Other cognitive measures

To demonstrate the specificity of the relationships of the above cognitive tests to degeneration in the SN and LC, we also examined performance in the domains of visuospatial function and language. Visuospatial function was assessed using the Clock Drawing Test (CDT) and the RCFT Copy trial. Language was assessed using the Letter Verbal Fluency (F, A, S; 1 minute per condition), Semantic Verbal Fluency (animals, actions; 1 minute per condition), and Boston Naming Test (errors without hints out of 60 items). The Montreal Cognitive Assessment (MoCA) was administered as a measure of global cognition.

### Neuroimaging

Participants underwent an MRI protocol which included 3D T1-weighted Magnetization-Prepared Rapid Acquisition Gradient Echo (T1w) and 2D T1-weighted Fast Spin Echo (neuromelanin-sensitive) sequences using a 3T Siemens Prisma scanner with a 32-channel head coil. Imaging parameters are listed in **Supplementary Table 2**. The present study aimed to investigate the structural integrity of the SN and LC using neuromelanin-sensitive images.

Raw neuromelanin-images underwent slice-by-slice normalization to remove inhomogeneities from the image using the MINC Toolkit ^63^ before being brought to stereotaxic space using transformations from each patient’s respective T1w image. Both sequences were acquired in the same session, with the assumption that they are co-registered in raw space. The raw T1w images had underwent pre-processing via the NeuroImaging & Surgical Technologies Longitudinal Pipeline^64^ and were non-linearly registered to stereotaxic space using the PD126 template^65^ as the registration target.

In standard space, the LC and SN regions of interest were defined using a conservatively thresholded probabilistic atlas,^66^ and expert manual segmentations on an in-house template, respectively. Neuromelanin signal intensity form these regions was calculated as a contrast-to-noise ratio, using the pontine tegmentum and cerebral peduncles as background intensity, respectively (1):

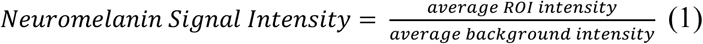

### Analyses

#### Reinforcement learning drift diffusion model

Reinforcement learning was evaluated by modelling dual learning rates.^30,48,67^ Computational modelling of the probabilistic learning task data was performed using the HDDM toolbox in Python.^68^ After removing omission errors from the dataset, we applied a reinforcement learning drift diffusion model (RLDDM), which uses the drift diffusion model as the choice policy (as opposed to typical softmax policies) within a reinforcement learning framework (i.e., Delta learning) allowing us to also account for cognitive processes related to decision-making.^69,70^ Evidence accumulation parameters were estimated using the drift diffusion model given that binary decision-making processes are required to engage in our reinforcement learning task, i.e., deciding between two stimuli on each trial, which provides more behavioural resolution than learning models using softmax or greedy choice policies that do not account for response time and thus speed-accuracy trade-offs.^71^

Briefly, in the RLDDM, learning is modelled using the delta learning rule with dual learning rates for positive and negative prediction errors, while the choice policy is modelled by the drift diffusion model. Drift rate (*v_i_*) is the speed of evidence accumulation, which is described as the per trial (*i*) difference between expected reward values between the two stimuli (*Q_upper, i_ – Q_lower, i_*) multiplied by a parameter (*v*) representing the degree of exploration or exploitation (analogous to the inverse temperature parameter in softmax choice policies). Decision boundary (*a*) is the distance between the two responses such that a greater distance represents greater response caution, i.e., slower but more accurate responses. Non-decision time (*t*) is the time associated with stimulus encoding and motor execution.

The RLDDM uses hierarchical and Bayesian estimation to fit the model. The hierarchical approach estimates an individual and group-level distribution of parameters. We specified group (i.e., Parkinson’s disease vs. Control) as a condition for each parameter such that each group had their respective distribution of parameters. There was an exception for non-decision time for which we estimated one group-level distribution spanning patients and controls together since this parameter does not represent a cognitive feature of behaviour. The Bayesian approach estimates a posterior distribution of parameter values using the Markov Chain Monte Carlo method. We generated 15,000 samples and discarded the first 5,000 samples. Convergence was assessed by visual inspection of the trace, autocorrelation, and histogram of the posteriors, and by removing participants with a Gelman-Rubin convergence diagnostic greater than 1.1.

Positive learning rate was our main parameter of interest as it represents the speed of updating value from positive feedback. Negative learning rate, drift rate and decision boundary served as control parameters; they measure different behavioural processes within the reinforcement learning task that were not expected to be related to dopamine neuron loss.

#### Statistical analyses

To evaluate group differences in demographic characteristics we conducted independent samples t-tests (two-tailed) for continuous, normally distributed variables, Mann-Whitney U tests for continuous, not normally distributed variables, and chi-squared analyses for categorical variables. To examine group differences in RLDDM-derived behavioural parameters on the reinforcement learning task, we computed a Bayesian p-value by calculating the proportion of each parameter’s posterior distribution that overlapped between groups. To examine group differences in neuromelanin signal intensities, we conducted ordinary least squares regressions with group as an independent variable (effect coded with Controls as the reference group) and controlled for age, sex, and years of education.

To investigate brain-behaviour relationships, we conducted ordinary least squares regressions on cognitive measures with SN and LC neuromelanin signal intensities as independent variables controlling for age, sex, and years of education. Reinforcement learning and decision-making behaviour was summarized by behavioural parameters from the RLDDM. Cognitive performance on the neuropsychological tests was summarized by five composite scores representing attention/working memory, executive function, memory, visuospatial function, and language. Performance in each neuropsychological task was z-scored (separately for Parkinson’s disease and Control groups) and domain composite scores were computed by averaging across z-scores that fell within respective domains. Each RLDDM-derived behavioural parameter and each cognitive composite score was evaluated in separate models as the dependent variable and separate analyses were conducted in Parkinson’s patients and Controls. We performed exploratory analyses when either the SN or LC neuromelanin signal intensity significantly predicted cognitive performance to investigate whether there was an interaction between them by repeating the analysis with the additional interaction term in the regression model. We also performed exploratory analyses decomposing the composite scores for attention/working memory, executive function, and declarative memory into their individual components to understand the relative contribution of each measure to the relationship with neuromelanin scores. To do this, we repeated the regressions replacing the dependent variable with each individual score.

For all regressions, sex was effect coded with males as the reference group and all continuous independent variables were z-scored (i.e. neuromelanin signal intensity scores, age and education), such that we could directly compare the magnitudes of the standardized beta estimates. Significance was determined by an alpha-level of less than 0.05.

## Results

### Reinforcement learning performance is impaired in PD

We compared reinforcement learning parameters between 117 participants with Parkinson’s disease (PD) and 65 Controls who completed the probabilistic learning task (**Supplementary Table 1**). As expected, PD participants had lower positive learning rates (*M_PD_* = 0.085, *M_CTRL_* = 0.227, *p* = 0.01) compared to Controls (**Fig. 2**). We also found lower drift rate in PD (*M_PD_* = 2.421, *M_CTRL_* = 3.598, *p* = 0.01) but no significant differences in negative learning rate (*M_PD_* = 0.005, *M_CTRL_* = 0.007, *p* = 0.345), or decision boundary (*M_PD_* = 1.922, *M_CTRL_* = 1.956, *p* = 0.282) relative to Controls.

**Figure 2.**
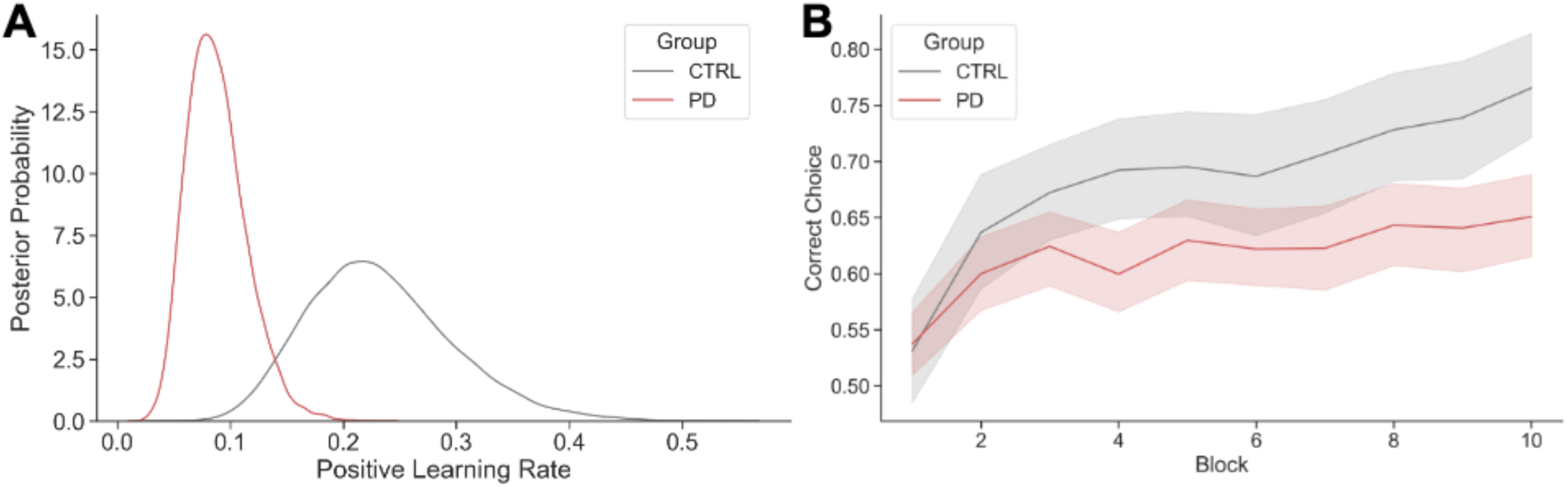
Reward learning differences between groups. **(A)** Posterior distributions of the positive learning rates demonstrate that participants with Parkinson’s disease (PD) are estimated to have lower positive learning rates compared to Controls (CTRL). **(B)** The raw data is depicted as an average of correct or incorrect optimal choices (0 or 1) across 10 blocks of 15 trials each.

### Cognitive performance on measures of LC degeneration is impaired in PD

A subset of 61 PD participants and 25 Control participants who completed all neuropsychological tests (for a total of 15 measures) were included in the computation of the domain composite scores (**Supplementary Table 1**). Overall, PD participants had numerically worse performance on all cognitive measures relating to attention/working memory, executive function and memory, but statistically significant differences were found on memory (the HVLT total and delayed) and executive function (TMT B-A, D-KEFS CWIT Inhibition) scores (**Table 2**). PD participants also had numerically worse performance on average in the tests of visuospatial function and language, but group differences were not statistically different.

**Table 2.**
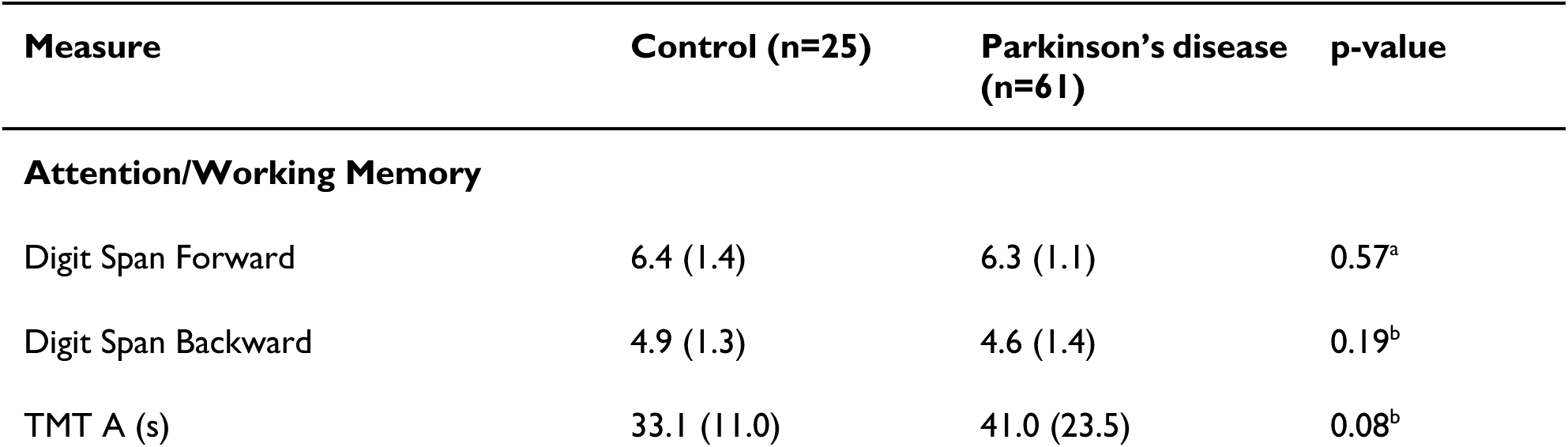
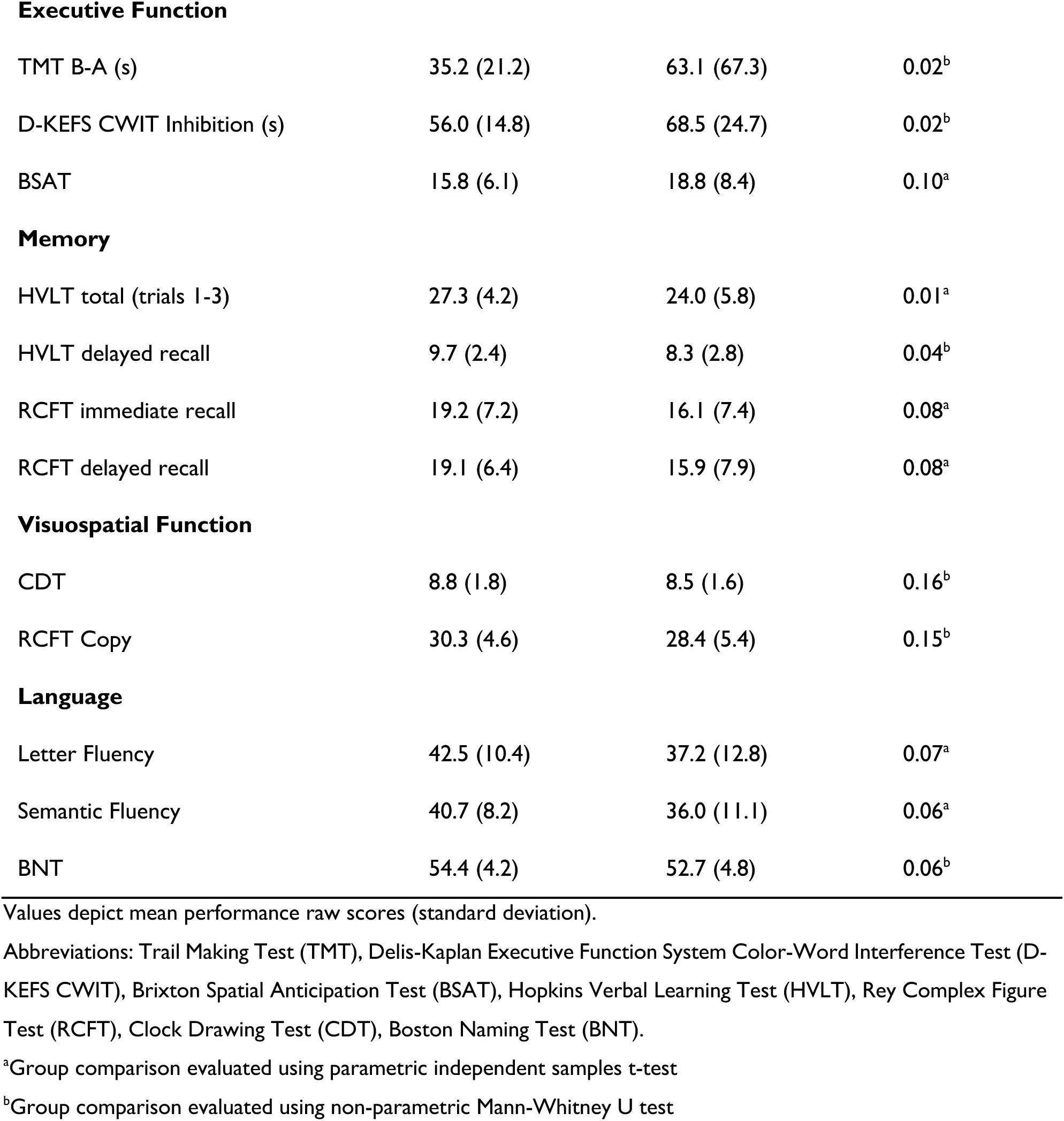
Group performance on neuropsychological assessments.

| Measure | Control (n=25) | Parkinson’s disease (n=61) | p-value |
| --- | --- | --- | --- |
| **Attention/Working Memory** |  |  |  |
| Digit Span Forward | 6.4 (1.4) | 6.3 (1.1) | 0.57<sup>a</sup> |
| Digit Span Backward | 4.9 (1.3) | 4.6 (1.4) | 0.19<sup>b</sup> |
| TMT A (s) | 33.1 (11.0) | 41.0 (23.5) | 0.08<sup>b</sup> |
| **Executive Function** |  |  |  |
| TMT B-A (s) | 35.2 (21.2) | 63.1 (67.3) | 0.02<sup>b</sup> |
| D-KEFS CWIT Inhibition (s) | 56.0 (14.8) | 68.5 (24.7) | 0.02<sup>b</sup> |
| BSAT | 15.8 (6.1) | 18.8 (8.4) | 0.10<sup>a</sup> |
| **Memory** |  |  |  |
| HVLT total (trials 1-3) | 27.3 (4.2) | 24.0 (5.8) | 0.01<sup>a</sup> |
| HVLT delayed recall | 9.7 (2.4) | 8.3 (2.8) | 0.04<sup>b</sup> |
| RCFT immediate recall | 19.2 (7.2) | 16.1 (7.4) | 0.08<sup>a</sup> |
| RCFT delayed recall | 19.1 (6.4) | 15.9 (7.9) | 0.08<sup>a</sup> |
| **Visuospatial Function** |  |  |  |
| CDT | 8.8 (1.8) | 8.5 (1.6) | 0.16<sup>b</sup> |
| RCFT Copy | 30.3 (4.6) | 28.4 (5.4) | 0.15<sup>b</sup> |
| **Language** |  |  |  |
| Letter Fluency | 42.5 (10.4) | 37.2 (12.8) | 0.07<sup>a</sup> |
| Semantic Fluency | 40.7 (8.2) | 36.0 (11.1) | 0.06<sup>a</sup> |
| BNT | 54.4 (4.2) | 52.7 (4.8) | 0.06<sup>b</sup> |

Values depict mean performance raw scores (standard deviation).
Abbreviations: Trail Making Test (TMT), Delis-Kaplan Executive Function System Color-Word Interference Test (D-KEFS CWIT), Brixton Spatial Anticipation Test (BSAT), Hopkins Verbal Learning Test (HVLT), Rey Complex Figure Test (RCFT), Clock Drawing Test (CDT), Boston Naming Test (BNT).
<sup>a</sup>Group comparison evaluated using parametric independent samples t-test
<sup>b</sup>Group comparison evaluated using non-parametric Mann-Whitney U test

### SN and LC neuromelanin signal intensities are reduced in PD

We investigated the group differences in neuromelanin signal intensities in 81 PD participants and 33 Controls who completed neuroimaging and the reinforcement learning task and/or all 15 neuropsychological measures to be able to relate MRI findings to cognitive performance. We found that PD participants had lower SN neuromelanin signal than Controls (*β_group_* = -0.87, *t* = - 4.46, *p* < 0.001, **Fig. 3**). There was also an effect of sex where men overall had lower SN neuromelanin signal than women (*β_sex_* = 0.46, *t* = 2.51, *p* = 0.01), but no effects of age or education (*β_age_* = 0.06, *t* = 0.78, *p* = 0.43; *β_education_* = 0.05, *t* = 0.65, *p* = 0.52). PD participants also had lower LC neuromelanin signal than Controls (*β_group_* = -0.52, *t* = -2.55, *p* = 0.01). Additionally, older age was associated with lower LC neuromelanin signal (*β_age_* = -0.27, *t* = -3.20, *p* = 0.002) but education and sex were not (*β_education_* = 0.05, *t* = 0.54, *p* = 0.59; *β_sex_* = 0.34, *t* = 1.78, *p* = 0.08). Overall, PD participants had evidence of greater SN and LC degeneration than Controls, with LC degeneration being additionally predicted by age.

**Figure 3.**
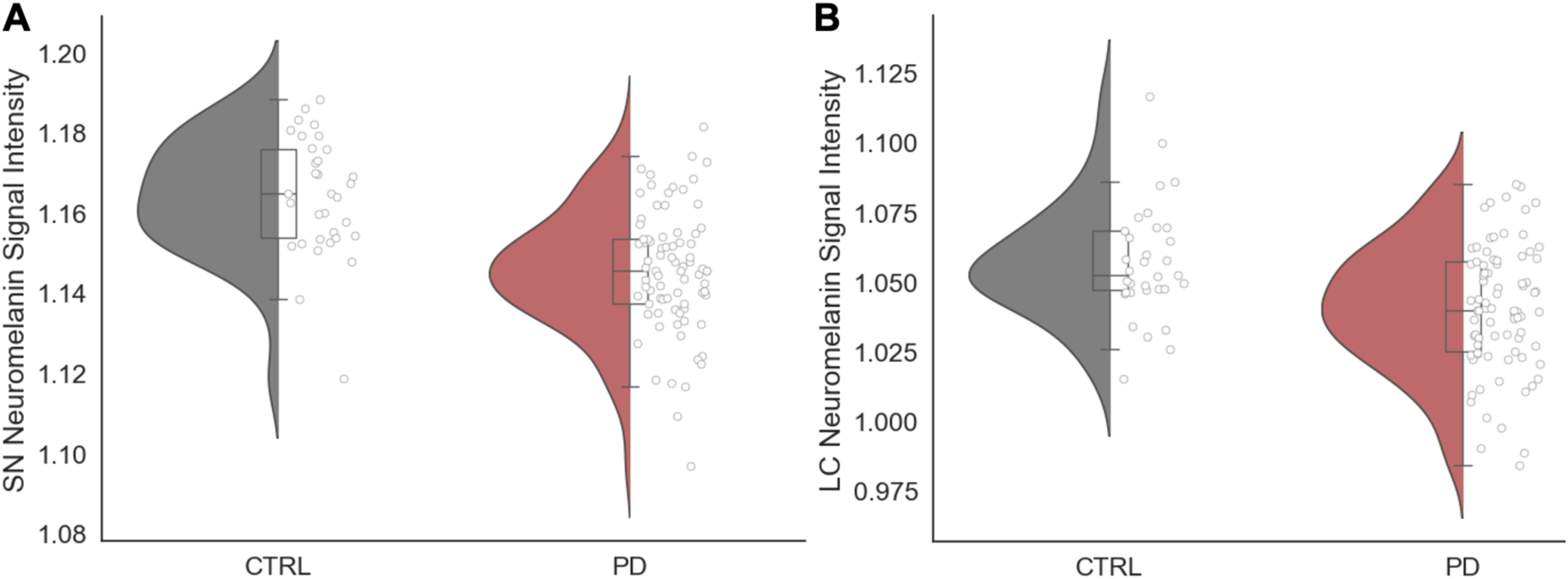
Neuromelanin signal intensity differences between groups. Participants with Parkinson’s disease (PD) have lower neuromelanin signal intensities compared to Controls (CTRL) in **(A)** the substantia nigra (SN) and **(B)** the locus coeruleus (LC). Box and whisker plots represent the median, interquartile interval, minimum, and maximum.

### Positive reinforcement learning is associated with SN degeneration

To understand the unique effect of SN degeneration in PD on reinforcement learning, we performed a linear regression on positive learning rate with both SN and LC neuromelanin signal intensities as independent variables and age, sex, and education as covariates. A subset of 62 PD participants and 25 Controls completed relevant measures and were included in the analysis (**Supplementary Table 1**). We found that in participants with PD, lower SN neuromelanin signal intensity was associated with lower positive learning rates (*β_SN_* = 0.41, *t* = 2.46, *p* = 0.02; **Fig. 4A**), but there was no association between LC neuromelanin and positive learning rate (*β_LC_* = 0.02, *t* = 0.14, *p* = 0.89; **Fig. 4A**). Age, sex, and education were not significant predictors of positive learning rate (*β_age_* = -0.08, *t* = -0.57, *p* = 0.57; *β_sex_* = -0.36, *t* = =1.04, *p* = 0.30; *β_education_* = -0.10, *t* = 0.72, *p* = 0.47). An exploratory analysis found no interaction between SN and LC signal intensity on positive learning rate *(β_SNxLC_* = -0.02, *t* = -0.09, *p* = 0.93, **Supplementary Table 5**). To demonstrate the selectivity of the relationship between SN neuromelanin and positive learning rate, we repeated analyses relating SN and LC neuromelanin to the other cognitive parameters derived from the RLDDM. SN and LC neuromelanin signal intensities were not associated with negative learning rate, drift rate, nor decision boundary (p > 0.29, **Supplementary Table 3**).

**Figure 4.**
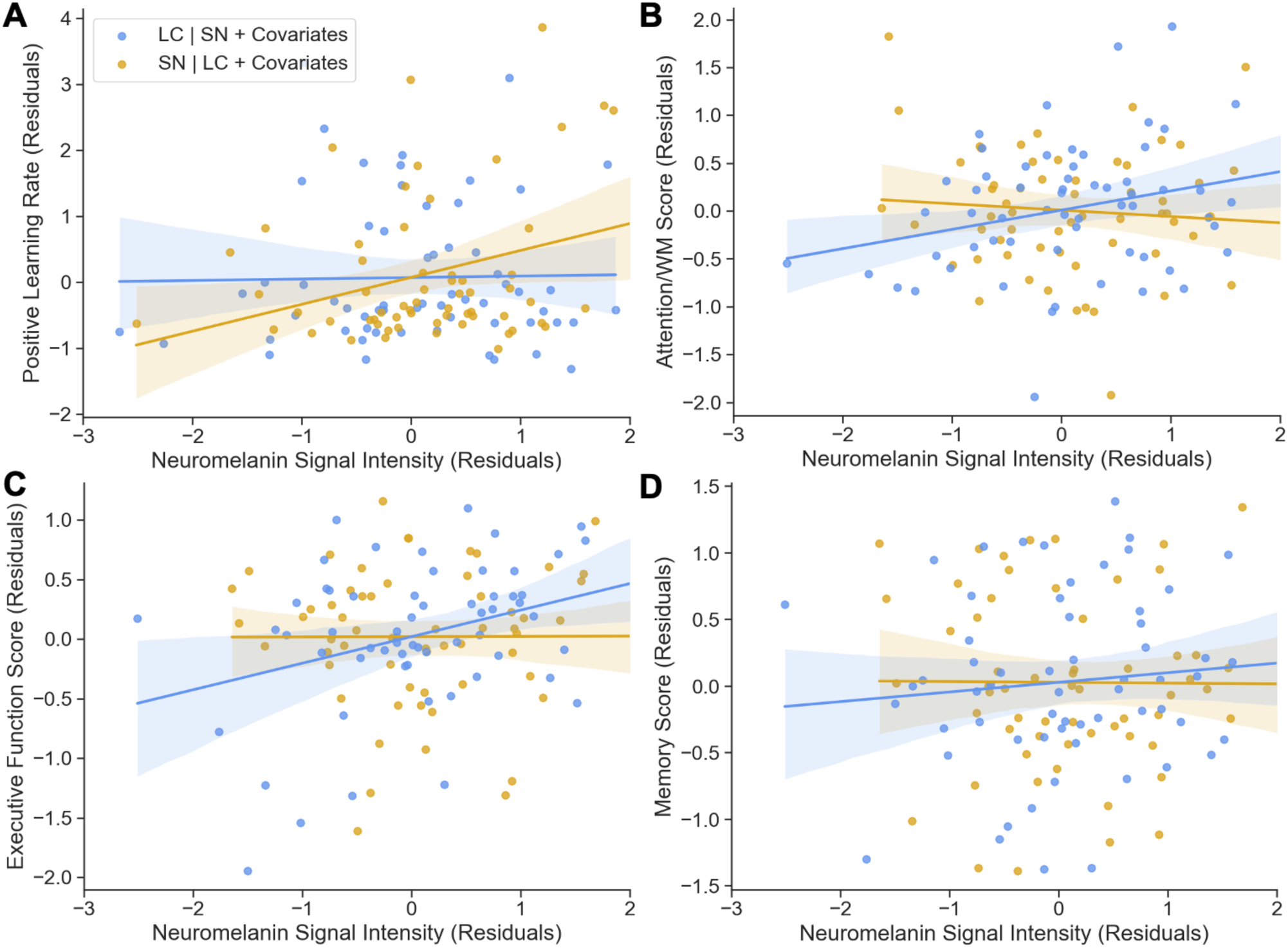
Brain-behaviour relationships in Parkinson’s participants. **(A)** Substantia nigra (SN), but not locus coeruleus (LC), signal intensity was associated with positive learning rate. LC, but not SN, signal intensity was associated with **(B)** attention/working memory (WM) and **(C)** executive functioning. **(D)** Neither SN nor LC signal intensity was associated with memory. Figures demonstrate the standardized beta coefficients after accounting for neuromelanin signal intensity in both the SN and LC, age, sex, and years of education. All variables are standardized.

To contextualize these results, we conducted an exploratory analysis in the smaller sample of Controls (*n* = 25). Surprisingly, higher SN neuromelanin signal intensity was associated with lower positive learning rates (*β_SN_* = -.49, *t* = -2.44, *p* = 0.02). There was no association between LC neuromelanin signal intensity and positive learning rate (*β_LC_* = -0.22, *t* = -1.16, *p* = 0.26; **Fig. 4A**) controlling for age, sex, and education (*β_age_* = 0.24, *t* = -1.16, *p* = 0.25; *β_sex_* = 0.35, *t* = 0.72, *p* = 0.48; *β_education_* = 0.26, *t* = 1.51, *p* = 0.15). However, the posterior distribution of positive learning rates in this smaller subsample of Controls was not representative of the posterior distribution of positive learning rates in the overall sample of Controls, thus this result should be interpreted with caution (**Supplementary** Fig. 1). Separate regressions also showed that SN and LC neuromelanin signal intensities were not associated with negative learning rate, drift rate, or decision boundary in Controls (p > 0.24, **Supplementary Table 3**).

In summary these results demonstrate that greater SN degeneration was selectively associated with worse positive reinforcement learning in PD participants, and that, importantly, there is no significant association of age with reinforcement learning.

### Attention/working memory and executive function are associated with LC degeneration

Next, we examined the unique relationship between LC neurodegeneration and performance in the three domains of cognition hypothesized to be related to LC in 61 PD participants and 25 Controls who completed all neuropsychological tests and neuroimaging. Models controlled for SN degeneration, in addition to age, sex and education, as above. In PD participants, as predicted, lower LC neuromelanin signal intensity was associated with worse attention/working memory performance (*β_LC_* = 0.20, *t* = 2.11, *p* = 0.01, **Fig. 4B**) and worse executive function performance (*β_LC_* = 0.22, *t* = 2.64, *p* = 0.01, **Fig. 4C**), but not with memory (*β_LC_* = 0.04, *t* = 0.41, *p* = 0.69, **Fig. 4D**). SN neuromelanin was not associated with performance in these three domains (attention/working memory: *β_SN_* = -0.07, *t* = -0.68, *p* = 0.50; executive function: *β_SN_* = 0.001, *t* = 0.02, *p* = 0.99; declarative memory: *β_SN_* = 0.01, *t* = 0.06, *p* = 0.95).

To further confirm the selective effects of SN and LC degeneration on cognition we also examined their relationship to visuospatial function and language. Neither SN nor LC degeneration were associated with performance in these domains (visuospatial: *β_SN_* = -0.15, *t* = -1.36, *p* = 0.18; *β_LC_* = 0.09, *t* = 0.87, *p* = 0.39; language: *β_SN_* = 0.08, *t* = 0.81, *p* = 0.42; *β_LC_* = 0.17, *t* = 1.70, *p* = 0.10) performance in PD participants. Across all five domains we found that older age was a predictor of worse performance (attention/working memory: *β_age_* = -0.28, *t* = -3.04, *p* = 0.004; executive function: *β_age_* = -0.40, *t* = -4.92, *p* < 0.001; memory: *β_age_* = -0.47, *t* = -4.82, *p* < 0.001; visuospatial: *β_age_* = -0.38, *t* = -3.54, *p* = 0.001; language: *β_age_* = -0.31, *t* = -3.23, *p* = 0.002). In all models, the standardized beta estimate for the effect of age on cognitive performance was larger than that of the effect of LC degeneration on cognitive performance. There were no effects of sex nor education on any of the cognitive domains (p > 0.16, see **Supplementary Table 4** for these results). To contextualize these results, we repeated analyses in Control participants. Neither SN nor LC neuromelanin signal intensity were significant predictors of any cognitive domain (p > 0.14, **Supplementary Table 4**).

We also performed exploratory analyses in PD participants predicting performance on each individual neuropsychological measure that fell into a cognitive domain that we hypothesized would relate to LC integrity, i.e., attention/working memory, executive function, and declarative memory. We posited that perhaps the individual scores were not equal in their representation of cognitive performance within their respective domains as is assumed in our composite score. Lower LC signal intensity was associated with worse performance on the D-KEFS CWIT Inhibition condition (*β_LC_* = 0.23, *t* = 2.01, *p* = 0.05), whereas SN signal intensity did not predict performance (*β_SN_* = -0.08, *t* = -0.71, *p* = 0.48). However, neither SN or LC neuromelanin signal intensity had a significant effect on Digit Span Forward, Digit Span Backward, TMT A, TMT B-A, BSAT errors, HVLT total and delayed recall, or RCFT immediate and delayed recalls performance (p > 0.07, see **Supplementary Table 6**).

Given that LC, but not SN, signal intensity was associated with performance on attention/working memory and executive function tasks, we performed exploratory analyses investigating whether there was additionally an interaction between LC and SN signal intensity on cognitive performance. We did not find an interaction between LC and SN signal intensity on attention/working memory (*β_SNxLC_* = -0.03, *t* = -0.29, *p* = 0.77) nor or executive function (*β_SNxLC_* = -0.08, *t* = -0.83, *p* = 0.41; see **Supplementary Table 5**).

In summary, we found that LC degeneration was selectively associated with impaired attention/working memory and executive function, but not, memory. However, it is also interesting to note that older age was a stronger predictor of worse cognitive performance than the degree of LC degeneration. This was not the case for the relationship between SN degeneration and reinforcement learning where age was not significantly associated with performance.

### Sensitivity analyses

All above analyses assessing brain-behaviour relationships were repeated in the 42 PD participants who completed all assessments, i.e., the reinforcement learning task, the complete neuropsychological assessment, and neuroimaging (**Supplementary Table 1**). The associations between SN neuromelanin and positive reinforcement learning (*β_SN_* = 0.28, *t* = 1.52, *p* = 0.14) and between LC neuromelanin and attention/working memory (*β_LC_* = 0.21, *t* = 1.87, *p* = 0.07) and executive function (*β_LC_* = 0.16, *t* = 1.81, *p* = 0.08) were positive, as above, suggesting that greater loss of neuromelanin was associated with worse performance. However, these associations did not reach significance, possibly due to lack of power in the smaller sample size.

## Discussion

Ascending neuromodulatory systems originating in the SN and LC play a pivotal role in supporting cognitive functions like attention, learning, memory, and executive function. In Parkinson’s disease, both SN and LC degeneration occur early. In the present study, we investigated the independent contributions of degeneration in each of these systems to the early cognitive deficits observed in this population. We found that in a sample of patients tested on their usual dose of dopaminergic medications, loss of neuromelanin in the SN – a marker of degeneration in dopamine neurons – was selectively associated with impaired learning from positive feedback, whereas loss of neuromelanin in the LC – a marker of degeneration noradrenergic neurons – was selectively associated with impairments in attention/working memory and executive function. Neither positive reinforcement learning performance nor SN degeneration were associated with age, whereas both LC degeneration and performance on LC-associated cognitive functions were strongly predicted by age in people with Parkinson’s disease. These results extend findings from pharmacological studies by showing that the severity of neurodegeneration in the SN and LC can be used to predict individual cognitive phenotypes in Parkinson’s disease. Furthermore, these results suggest that the brain-behaviour relationship linking SN degeneration to positive reinforcement learning provides a window into the Parkinson’s-specific and age-independent effect of neurodegeneration on cognition. Given increasing recognition that age-associated pathologies are prevalent in Parkinson’s disease and contribute to cognitive outcomes, these results highlight the potential value of measuring brain-behaviour relationships that are informed by mechanisms to begin to parse the multiple causes of cognitive impairment that likely play a role in Parkinson’s disease.

### SN degeneration is selectively associated with impaired positive reinforcement learning

First, we found that the severity of SN degeneration, as measured by loss of neuromelanin signal, was associated with reduced learning from positive feedback in participants with Parkinson’s disease. This relationship is consistent with the extensive literature demonstrating that reduced dopamine signalling is associated with impaired positive reinforcement learning and that there is a beneficial effect of dopamine replacement therapy on performance, though not all studies show this.^30,48,51,53,72^ The current findings extend our current understanding of the role of dopamine depletion in cognitive impairment in a few important ways. First, rather than studying the effects of a fixed dose of dopamine replacement, which cannot provide insight into the severity of the underlying dopamine deficit, we used neuromelanin MRI, one of the most promising biomarkers of degeneration in Parkinson’s disease, to establish the severity of degeneration.^73–76^ Our results therefore suggest that positive reinforcement learning could be used as a behavioural performance-based assessment to track SN degeneration, complimentary to the way motor assessments also correlate with neuromelanin signal.^1,40,74,77,78^ Second, the relationship between SN neuromelanin signal and positive reinforcement learning was independent of age, and, perhaps even more critical, age was not a predictor of the computationally-derived measure of learning performance we used. This suggests that positive reinforcement learning performance could be useful as a performance-based metric to track the specific Parkinson’s-related, rather that age-related, effects of neurodegeneration.

Of note is that positive reinforcement learning performance was impaired in Parkinson’s participants even though they were on dopaminergic medication at the time of testing. Several previous studies, but not all,^49,59,79,80^ have shown some degree of remediation of learning deficits in the medicated state.^30,31,48–53^ One possible explanation for the presence of a learning impairment in the medicated Parkinson’s participants in our sample is that they have somewhat more advanced disease than the typical samples of participants recruited to medication manipulation studies, where the requirement of completing an OFF medications session necessarily biases the sample towards less affected participants. Indeed, this is also consistent with the fact that, apart from patients with very mild disease, dopamine replacement rarely completely remediates the dopamine-sensitive motor symptoms and therefore should not be expected to completely remediate any dopamine-sensitive cognitive symptoms. Demonstrating that reinforcement learning performance captures individual differences in SN neurodegeneration even in medicated patients is a strength as it suggests this measure could be useful in the clinical research setting. Future research could additionally include a measure of the total dose of dopaminergic medications in the models to account for possible confounding effects and obtain a more precise measurement of the relationship between SN degeneration and positive reinforcement learning.

There was no relationship between SN neuromelanin and any other cognitive performance measure, including the other computationally derived measures from the reinforcement learning task. This is largely in keeping with the few other studies that have investigated the relationship between SN degeneration and cognitive performance in participants with Parkinson’s disease, though some results have been inconsistent.^1,39,40^ For instance, one study found a relationship between SN integrity and working memory^39^ and another study found a relationship between anteromedial and superior SN integrity and attention.^40^ These contradictory results may be explained by different working memory and attention tasks used across studies which may be differentially sensitive to deficits. In particular, the Digit Span task we used did not differentiate groups and has not been shown to be sensitive to dopamine replacement, raising questions about whether a relationship to SN integrity is to be expected.^81,82^ However, most notably, given the strong relationship between LC degeneration and cognitive performance, prior studies have not controlled for the jointly occurring LC degeneration in analyses of the impact of SN degeneration on cognition.

### LC degeneration is selectively associated with attention, working memory and executive function

We found that LC degeneration was associated with impaired attention and executive function in Parkinson’s disease participants, but not with declarative memory, visuospatial, language, nor positive reinforcement learning. This is largely consistent with previous studies^1,2,5,54,55^ and has been proposed to reflect the fact that frontal cortical regions like the prefrontal cortex (PFC) – that are thought to play a critical role in top-down attention, working memory, and executive function^83–87^ – are also projection sites of LC neurons,^88^ suggesting that the neural mechanism by which Parkinson’s disease participants experience these impairments may be, at least in part, via the loss of innervation from the degenerating LC to the PFC. We did not replicate previously reported relationships between LC degeneration and declarative memory deficits, though this has primarily been observed in older adults.^1,3,4,89,90^ While this could reflect the fact that our memory measures may not have been sufficiently selective (for instance, the RCFT is a measure of visual memory that also engages visuospatial function), this also raises the possibility that memory deficits in Parkinson’s disease primarily reflect involvement of other structures – such as the hippocampus, amygdala, or nucleus basalis of Meynert.^91–93^

### Limitations

An important limitation of our study was that we did not obtain all measurements in all participants, which limited our ability to examine interactions between both regions of interest and their related domains of cognition. In particular, the sample of older adult controls with complete testing was small thereby limiting our ability to perform meaningful contrasts and to fully account for the effects of age in the Parkinson’s disease participants. The effect of age is especially difficult to estimate in Parkinson’s disease given its relationship to disease duration but is critical to consider because age-associated brain changes also occur in Parkinson’s disease and appear to contribute to the cognitive phenotype in ways that overlap with the effects of neurodegeneration. A second limitation is that we had only one positive reinforcement learning measure. Given that the literature examining dopaminergic medication effects on positive reinforcement learning has revealed conflicting results that might depend on the type of task used, it will be important to replicate the association we found between SN degeneration and positive reinforcement learning. This would also be an opportunity to develop a shorter task, which would be more practical for implementation in clinical studies.

### Summary

The goal of this study was to identify aspects of the cognitive phenotype of Parkinson’s disease that can be selectively attributed to SN and LC degeneration. We found that SN degeneration was selectively associated with positive reinforcement learning impairments and that LC degeneration was selectively associated with impairments in attention and executive function. Identifying dissociable roles of SN and LC degeneration on different cognitive functions in early Parkinson’s disease that reflect the effects of disease above and beyond the effects of age has important implications for developing performance-based measures of cognitive function in Parkinson’s disease that specifically reflect Parkinson’s-related neurodegeneration. These findings also pave the way towards using neurobiologically-grounded performance-based measures of brain function to begin to identify different sources underlying the heterogeneity in the cognitive and behavioural phenotype of Parkinson’s disease.

## Data availability

De-identified participant data is available upon request.

## Acknowledgements

The authors would like to thank the Quebec Parkinson Network and all participants without whom this study would not be possible.

