## Supplementary Materials for "Selective effects of dopaminergic and noradrenergic degeneration on cognition in Parkinson’s disease"

**Supplementary Table 1.** Characteristics for each subsample

| Subsample | Completed PLT | | | Completed Imaging | | | Completed PLT + Imaging | | | Completed Neuropsych + Imaging | | | Completed PLT + Neuropsych + Imaging | | |
|---|---|---|---|---|---|---|---|---|---|---|---|---|---|---|---|
| | CTRL (n=65) | PD (n=117) | p-value | CTRL (n=33) | PD (n=81) | p-value | CTRL (n=25) | PD (n=62) | p-value | CTRL (n=25) | PD (n=61) | p-value | CTRL (n=17) | PD (n=42) | p-value |
| Age[a] | 64.0 (9.7) | 64.4 (8.9) | 0.81 | 62.5 (10.7) | 63.5 (9.9) | 0.64 | 63.6 (10.2) | 63.8 (9.5) | 0.92 | 62.1 (10.9) | 64.8 (9.9) | 0.27 | 63.3 (10.5) | 65.2 (9.9) | 0.50 |
| Males/Females[b] | 24/41 | 83/35 | <0.001 | 9/24 | 57/24 | <0.001 | 6/19 | 43/19 | <0.001 | 5/20 | 43/18 | <0.001 | 3/14 | 29/13 | <0.001 |
| Education, years[a] | 15.7 (3.1) | 15.1 (3.2) | 0.21 | 15.8 (3.1) | 14.5 (3.7) | 0.09 | 15.8 (3.1) | 14.6 (3.5) | 0.15 | 15.5 (3.8) | 14.3 (3.8) | 0.17 | 16.5 (3.2) | 14.1 (3.8) | 0.02 |
| MoCA[c] | 27.3 (1.8) | 25.8 (2.8) | 0.01 | 27.2 (2.2) | 25.1 (3.1) | 0.02 | 27.4 (1.9) | 25.7 (2.7) | 0.01 | 27.3 (2.2) | 25.4 (3.1) | 0.04 | 27.5 (1.9) | 25.5 (2.8) | 0.05 |
| Disease Duration | | 5.0 (4.3) | | | 5.5 (4.5) | | | 5.7 (4.9) | | | 5.2 (4.3) | | | 5.7 (4.9) | |
| UPDRS-III | | 30.0 (13.0) | | | 26.2 (14.1) | | | 28.9 (13.7) | | | 26.2 (14.3) | | | 28.0 (14.6) | |
| Questionnaire for Impulsive-Compulsive Disorders in Parkinson's disease[c] | 15.0 (10.7) | 17.1 (10.6) | 0.19 | 11.5 (7.1) | 17.7 (10.7) | 0.04 | 11.7 (7.1) | 17.6 (10.9) | 0.07 | 12.9 (17.1) | 17.1 (8.4) | 0.15 | 12.9 (17.1) | 17.1 (8.4) | 0.15 |
| Geriatric Depression Scale[c] | 1.7 (3.0) | 3.9 (4.1) | <0.001 | 1.1 (1.5) | 3.5 (4.0) | 0.03 | 1.2 (1.5) | 3.6 (4.1) | 0.04 | 1.2 (1.6) | 3.1 (3.8) | 0.17 | 1.2 (1.6) | 3.1 (3.8) | 0.17 |

Values depict the mean (standard deviation).
Abbreviations: Probabilistic Learning Task (PLT), Control (CTRL), Parkinson's disease (PD), Montreal Cognitive Assessment (MoCA), Unified Parkinson's Disease Rating Scale (UPDRS),.
The number of participants who completed the mood and behavioural questionnaires range from 43-44 CTRL & 80-86 PD for the PLT sample, 19 CTRL & 43-47 PD for the Imaging sample, 18 CTRL & 41-44 PD for the PLT + Imaging sample, 13 CTRL & 27-29 PD for the Neuropsych + Imaging and PLT + Neuropsych + Imaging samples
[a]Group comparison evaluated using parametric independent samples t-test
[b]Group comparison evaluated using chi-square test
[c]Group comparison evaluated using non-parametric Mann-Whitney U test

**Supplementary Table 2.** MRI sequence parameters

| MRI sequence name | Dimension | Relaxation time, TR | Echo time, TE | Flip Angle | Field of view for 2D acquisitions | Slice thickness | 3D volumetric resolution | Slice orientation | Matrix size (xyz) | Voxel size |
|---|---|---|---|---|---|---|---|---|---|---|
| 3D T1 Magnetization Prepared – Rapid Gradient Echo (MPRAGE) | 3D | 2300.0 ms | 2.98 ms | 9° | NA | 1.0 mm | 192 x 256 x 256 $mm^3$ | Sagittal | 192 x 256 x 256 | 1 x 1 x 1 $mm^3$ |
| T1-weighted Fast Spin Echo (neuromelanin-sensitive) | 2D | 600.0 ms | 10 ms | 120° | 165 x 220 $mm^2$ | 1.8 mm | NA | T > C4.9 | 240 x 320 x 20 | 0.7 x 0.7 x 1.8 $mm^3$ |

**Supplementary Table 3.** Standardized beta estimates (standard error) from linear regressions predicting learning rates and decision-making parameters

| Dependent Variable | Intercept (SE) | SN (SE) | LC (SE) | Age (SE) | Education (SE) | Sex (SE) |
|---|---|---|---|---|---|---|
| *Parkinson's patients (N=62)* | | | | | | |
| Positive learning rate | 0.09 (0.16) | 0.41 (0.17)* | 0.02 (0.15) | -0.08 (0.14) | 0.10 (0.13) | -0.36 (0.34) |
| Negative learning rate | -0.22 (0.04) | -0.005 (0.05) | -0.04 (0.04) | -0.01(0.04) | 0.01 (0.04) | -0.08 (0.09) |
| Drift rate | 0.03 (0.15) | 0.03 (0.15) | 0.11 (0.14) | -0.16 (0.13) | 0.05 (0.12) | 0.33 (0.32) |
| Decision threshold | -0.15 (0.14) | -0.15 (0.14) | -0.02 (0.13) | -0.16 (0.12) | -0.22 (0.11) | 0.23 (0.28) |
| *Controls (N=25)* | | | | | | |
| Positive learning rate | -0.15 (0.21) | -0.49 (0.20)* | -0.23 (0.19) | 0.24 (0.21) | 0.26 (0.18) | 0.35 (0.49) |
| Negative learning rate | -0.04 (0.41) | -0.49 (0.39) | -0.18 (0.38) | 0.04 (0.40) | 0.14 (0.34) | 0.84 (0.96) |
| Drift rate | 0.30 (0.29) | -0.004 (0.27) | -0.15 (0.26) | -0.63 (0.28) | 0.01 (0.24) | -1.08 (0.67) |
| Decision threshold | -0.11 (0.25) | -0.17 (0.24) | -0.34 (0.23) | -0.26 (0.25) | 0.54 (0.21) | -0.0003 (0.59) |

* $p < 0.05$; ** $p < 0.01$; *** $p < 0.001$

**Supplementary Table 4.** Standardized beta estimates (standard error) from linear regressions predicting cognitive domains

| Dependent Variable | Intercept (SE) | SN (SE) | LC (SE) | Age (SE) | Education (SE) | Sex (SE) |
|---|---|---|---|---|---|---|
| *Parkinson's patients (N=61)* | | | | | | |
| Attention & working memory | 0.01 (0.10) | -0.07 (0.10) | 0.20 (0.10)* | -0.28 (0.09)** | 0.07 (0.09) | 0.05 (0.22) |
| Executive function | 0.02 (0.09) | 0.001 (0.09) | 0.22 (0.08)* | -0.40 (0.08)*** | 0.003 (0.08) | 0.12 (0.20) |
| Memory | 0.01 (0.10) | 0.01 (0.10) | 0.04 (0.10) | -0.47 (0.10)*** | 0.09 (0.10) | 0.09 (0.10) |
| Visuospatial function | 0.04 (0.11) | -0.15 (0.11) | 0.09 (0.11) | -0.38 (0.11)** | 0.08 (0.11) | 0.18 (0.26) |
| Language | 0.07 (0.10) | 0.08 (0.10) | 0.17 (0.10) | -0.31 (0.10)** | 0.02 (0.10) | 0.32 (0.23) |
| *Controls (N=25)* | | | | | | |
| Attention & working memory | 0.30 (0.17) | -0.10 (0.14) | -0.02 (0.19) | -0.55 (0.18)** | 0.14 (0.15) | -0.98 (0.36)* |
| Executive function | 0.05 (0.17) | -0.02 (0.14) | 0.30 (0.19) | -0.16 (0.19) | 0.18 (0.15) | -0.15 (0.37) |
| Memory | -0.03 (0.22) | 0.01 (0.18) | 0.05 (0.24) | -0.10 (0.23) | 0.35 (0.19) | 0.11 (0.46) |
| Visuospatial function | 0.01 (0.19) | 0.04 (0.16) | -0.02 (0.21) | -0.46 (0.20)* | 0.63 (0.17)** | -0.04 (0.40) |
| Language | 0.06 (0.21) | -0.003 (0.17) | -0.05 (0.23) | -0.18 (0.22) | 0.40 (0.18)* | -0.19 (0.44) |

* $p < 0.05$; ** $p < 0.01$; *** $p < 0.001$

**Supplementary Table 5.** Standardized beta estimates (standard error) from exploratory regressions investigating SN and LC interaction

| Dependent Variable | Intercept (SE) | SN (SE) | LC (SE) | SN*LC (SE) | Age (SE) | Education (SE) | Sex (SE) |
|---|---|---|---|---|---|---|---|
| Positive learning rate | 0.09 (0.17) | 0.40 (0.17)* | 0.02. (0.16) | -0.02 (0.19) | -0.08 (0.14) | 0.10 (0.14) | -0.35 (0.25) |
| Attention & working memory | 0.02 (0.10) | -0.07 (0.10) | 0.20 (0.10)* | -0.03 (0.11) | -0.29 (0.09)** | 0.07 (0.10) | 0.05 (0.22) |
| Executive function | 0.04 (0.09) | 0.003 (0.09) | 0.21 (0.09)* | -0.08 (0.09) | -0.41 (0.08)*** | 0.01 (0.08) | 0.12 (0.20) |

* $p < 0.05$; ** $p < 0.01$; *** $p < 0.001$

**Supplementary Table 6.** Standardized beta estimates (standard error) from linear regressions predicting neuropsychological performance from the Attention/Working Memory, Executive Function, and Memory cognitive domains

| Dependent Variable | Intercept (SE) | SN (SE) | LC (SE) | Age (SE) | Education (SE) | Sex (SE) |
|---|---|---|---|---|---|---|
| Digit Span Forward | 0.03 (0.14) | -0.12 (0.14) | 0.19 (0.14) | -0.20 (0.14) | -0.05 (0.14) | 0.12 (0.33) |
| Digit Span Backward | -0.03 (0.14) | -0.03 (0.14) | 0.20 (0.14) | -0.22 (0.14) | 0.09 (0.14) | -0.18 (0.32) |
| TMT-A (s) | 0.04 (0.12) | -0.06 (0.12) | 0.21 (0.12) | -0.43 (0.12)** | 0.17 (0.12) | 0.22 (0.28) |
| TMT-B-A (s) | 0.07 (0.13) | -0.05 (0.13) | 0.24 (0.13) | -0.32 (0.13)* | -0.18 (0.13) | 0.32 (0.30) |
| D-KEFS CWIT Inhibition (s) | 0.11 (0.12) | -0.08 (0.12) | 0.23 (0.11)* | -0.45 (0.11)*** | -0.02 (0.11) | 0.53 (0.27) |
| BSAT | -0.10 (0.12) | 0.13 (0.12) | 0.20 (0.12) | -0.44 (0.12)*** | 0.21 (0.12) | -0.49 (0.28) |
| HVLT Total (Trials 1-3) | 0.12 (0.13) | 0.01 (0.13) | -0.01 (0.12) | -0.47 (0.12)*** | 0.02 (0.12) | 0.56 (0.29)* |
| HVLT Delayed | 0.10 (0.13) | -0.05 (0.13) | 0.17 (0.13) | -0.41 (0.12)* | -0.09 (0.12) | 0.49 (0.29) |
| RCFT Immediate | -0.04 (0.13) | -0.02 (0.13) | 0.06 (0.13) | -0.47 (0.12)*** | 0.12 (0.12) | -0.17 (0.30) |
| RCFT Delayed | -0.04 (0.13) | 0.02 (0.13) | 0.08 (0.13) | -0.47 (0.12)*** | 0.14 (0.12) | -0.18 (0.29) |

* $p < 0.05$; ** $p < 0.01$; *** $p < 0.001$

**Supplementary Table 7.** Confirmatory regressions in a subsample of 42 PD patients

| Dependent Variable | Intercept (SE) | SN (SE) | LC (SE) | Age (SE) | Education (SE) | Sex (SE) |
|---|---|---|---|---|---|---|
| Positive learning rate | -0.01 (0.17) | 0.28 (0.19) | -0.02 (0.17) | -0.26 (0.16) | -0.04 (0.17) | -0.06 (0.40) |
| Attention & working memory | 0.02 (0.12) | -0.21 (0.12) | 0.21 (0.11) | -0.33 (0.11)** | -0.15 (0.11) | 0.40 (0.27) |
| Executive function | 0.09 (0.09) | -0.07 (0.10) | 0.16 (0.09) | -0.43 (0.08)*** | -0.02 (0.09) | 0.25 (0.21) |

* $p < 0.05$; ** $p < 0.01$; *** $p < 0.001$

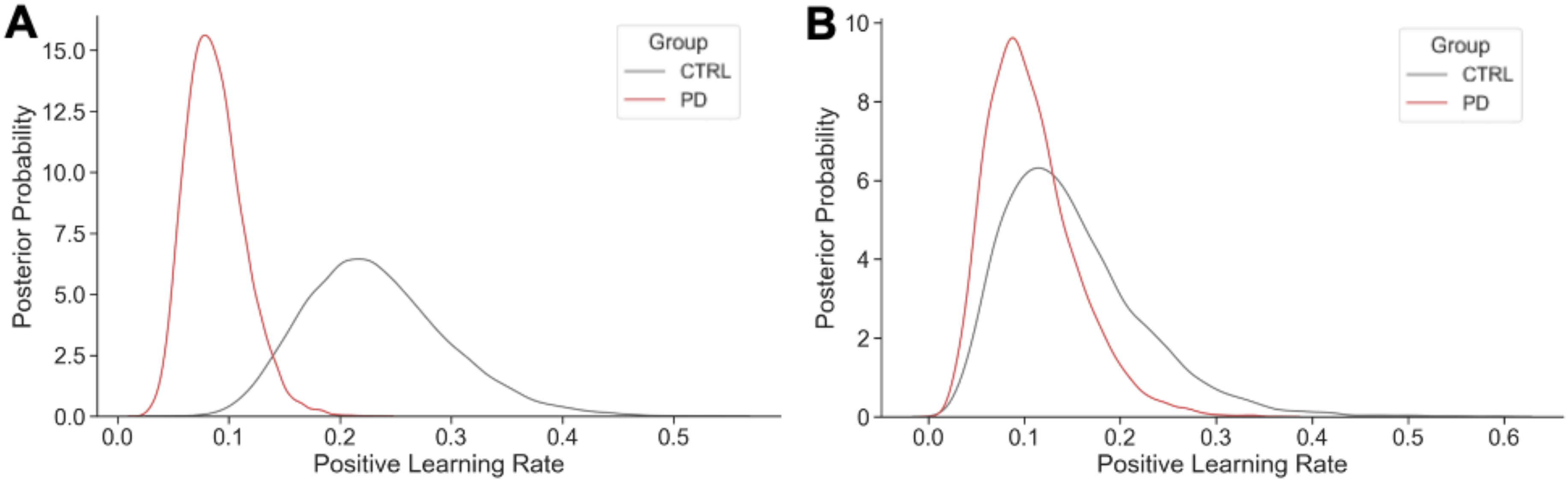


**Supplementary Figure 1. (A)** Posterior distribution of positive learning rates in 117 participants with Parkinson's disease (PD) and 65 Controls (CTRL) who completed the reinforcement learning task. PD participants are estimated to have lower positive learning rates than CTRLs (p=0.01). **(B)** Posterior distribution of positive learning rates in 62 participants with PD and 25 CTRLs who completed the reinforcement learning task *and* neuroimaging. In this smaller sample of participants, there are no group differences in positive learning rate (p=0.31). The smaller sample of PD participants have a similar distribution as the larger sample of PD participants, whereas the smaller sample of CTRLs have lower positive learning rates than the larger sample of CTRLs.
